# The hospital sink drain biofilm resistome is independent of the corresponding microbiota, the environment and disinfection measures

**DOI:** 10.1101/2024.12.07.24318639

**Authors:** Aurélie Hennebique, Jostin Monge-Ruiz, Morgane Roger-Margueritat, Patrice Morand, Claire Terreaux-Masson, Max Maurin, Corinne Mercier, Caroline Landelle, Elena Buelow

**Author notes:** these authors contributed equally (Aurélie Hennebique, Jostin Monge-Ruiz). BIP (Bioénergétique et Ingénierie des Protéines), CNRS/Aix-Marseille Université, Marseille, France.

## Abstract

In hospitals, the transmission of antibiotic-resistant bacteria (ARB) may occur via biofilms present in sink drains, which can lead to infections. Despite the potential role of sink drains in the transmission of ARB in nosocomial infections, routine surveillance of these drains is lacking in most hospitals. As a result, there is currently no comprehensive understanding of the transmission of ARB and the dissemination of antimicrobial resistance genes (ARGs) and associated mobile genetic elements (MGEs) via sink drains. This study employed a multifaceted approach to monitor the total aerobic bacterial flora as well as the presence of carbapenemase-producing *Enterobacterales* (CPEs), the microbiota and the resistome of sink drain biofilms (SDBs) and hospital wastewater (WW) of two separate intensive care units (ICUs) in the same healthcare facility in France. Samples of SDB and WW were collected on a monthly basis, from January to April 2023, in the neonatal (NICU) and the adult (AICU) ICUs of Grenoble Alpes University Hospital. In the NICU, sink drain disinfection with surfactants was performed routinely. In the AICU, however, routine disinfection is not carried out. No CPEs were isolated from SDBs in either ICU by bacterial culture. Culture-independent approaches revealed an overall distinct microbiota composition of the SDBs in the two units. The AICU SDBs were dominated by potential Gram-negative bacterial pathogens including *Pseudomonas*, *Stenotrophomonas*, *Staphylococcus*, and *Klebsiella*, while the NICU SDBs were dominated by the Gram-negative genera *Achromobacter*, *Serratia*, and *Acidovorax*, as well as the Gram-positive genera *Weisella* and *Lactiplantibacillus*. In contrast, the resistome of the SDBs exhibited no significant differences between the two units, indicating that the abundance of ARGs and MGEs is independent of microbiota composition and disinfection practices. Investigation of the WW that connected to the respective ICUs revealed that the WW from the AICU exhibited a more diverse aerobic flora than the WW from the NICU. In addition, the AICU WW yielded 15 CPEs, whereas the NICU WW yielded a single CPE. The microbiota of the NICU and AICU WW samples differed from their respective SDBs and exhibited distinct variations over the four-month period. It is noteworthy that the WW from the AICU contained a greater number of genes conferring resistance to quinolones and integron integrase genes, whereas the NICU exhibited a higher abundance of streptogramin resistance genes. The results of our study demonstrated that the resistome of the hospital SDBs in the two ICUs of the investigated healthcare institute is independent of the microbiota, the environment, and the local disinfection measures. However, a difference was observed in the prevalence of CPEs in the WW pipes collecting the waste from the investigated drains. These findings offer valuable insights into the resilience of resistance genes in SDBs in ICUs, underscoring the necessity for innovative strategies to combat antimicrobial resistance in clinical environments.

**Graphical abstract:** 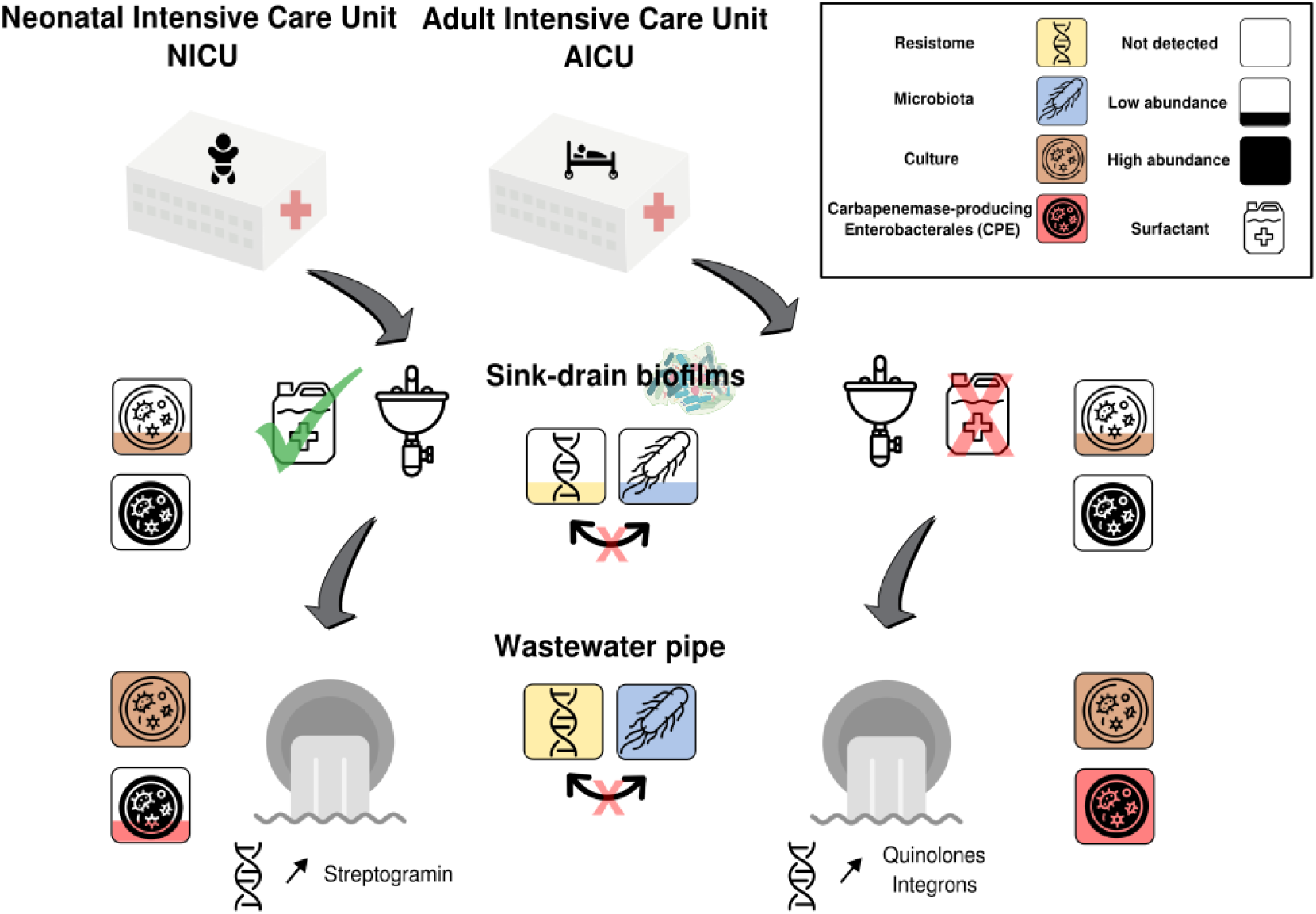

## 1. Introduction

In hospital settings, patient infections can occur via transmission of antibiotic resistant bacteria (ARB) from bacterial biofilms colonizing sink drains in patient rooms (De Geyter et al., 2021; Suleyman et al., 2018; van der Zwet et al., 2022). Sink drain biofilms (SDBs) can harbor nosocomial pathogens as well as highly resistant pathogens such as carbapenemase-producing *Enterobacterales* (CPEs), and glycopeptide-resistant Enterococci (GRE), which pose a significant threat to patients hospitalized in intensive care units (ICUs). The incidence of hospital-acquired infections linked to sink drains was shown to be reduced through the implementation of continuous or sporadic disinfection measures (Bourdin et al., 2024; Ledwoch et al., 2020) and was further shown to be significantly diminished in water-free healthcare settings (Hopman et al., 2017; Low et al., 2024). As a consequence, strict preventive measures in ICUs in some hospitals including screening and routine sink drain disinfection are implemented (Bourdin et al., 2024; de Jonge et al., 2019; van der Zwet et al., 2022). However, there is no consensus on the most effective strategies for controlling transmission of ARBs from sink drains to patients. Furthermore, the impact of disinfectants on the distribution and prevalence of microbial species (the microbiota), ARB, antibiotic resistance genes (ARGs), and associated mobile genetic elements (MGEs) in hospital sink drains and SDBs remains understudied (Catho et al., 2024; Ramos-Castaneda et al., 2020; Volling et al., 2021).

Biofilms are complex communities of microorganisms embedded in a self-produced extracellular matrix that facilitates their attachment to various surfaces, including sink drains and wastewater (WW) systems (Ledwoch et al., 2020; Maillard and Centeleghe, 2023; Wingender and Flemming, 2011). Biofilms are recognized as potential reservoirs of ARB and bacterial pathogens, which poses significant public health concerns (Balcazar et al., 2015; Maillard and Centeleghe, 2023). The extracellular matrix of biofilms provides protection against environmental stresses, including antibiotics and disinfectants, which makes eradication of biofilms challenging (Ledwoch et al., 2020; Wingender and Flemming, 2011). Biofilms formed in sink drains have shown particular resistance to disinfection interventions (Bourdin et al., 2024; Ledwoch et al., 2020; Maillard and Centeleghe, 2023; Pirzadian et al., 2020). Biofilms are important hotspots for horizontal gene transfer (HGT) (Abe et al., 2020) favoring the potential emergence of ARB in SDBs. A high prevalence of ARB, pathogens, ARGs, and MGEs was also described in hospital WW systems that collect global WW from hospital buildings, including sink drains (Buelow et al., 2023, 2020, 2018; Rodriguez-Mozaz et al., 2015). Moreover, WW and WW biofilms (WWB) act as breeding grounds for the emergence of ARB and the dissemination of ARGs *via* MGEs in the wider environment (Buelow et al., 2023, 2020, 2018). However, to which degree hospital sink drains and their biofilm populations impact the hospital WW composition remains unexplored, and whether hospital WW reflects the SD and SDB population remains unclear. Furthermore, the dynamics of the SDB microbiota and their resistome over time, particularly in response to hospital disinfection measures, remain largely unexplored.

A recent *in vitro* SDB model demonstrated the differential impact of various biocidal products on SDB microbiota, revealing reductions in bacterial counts limited to ∼ four days. This underscores the need for continuous disinfection (Ledwoch et al., 2020). Nevertheless, the prevalence and the abundance of ARGs and MGEs have yet to be assessed in this context. A comprehensive understanding of the dynamics of the SDB resistome is crucial to assess accurately its potential role in the emergence of ARB and the efficacy of disinfection interventions.

In the neonatal ICU (NICU) of the Grenoble Alpes University Hospital, sink drain disinfection with surfactants is conducted on a regular basis. In contrast, the adult ICU (AICU) has not implemented routine disinfection. At present, there is no routine monitoring of SDBs or hospital WW for the presence of CPEs. Furthermore, the dynamics of the microbiota and resistome of SDBs in ICUs and their respective WW remain poorly understood.

The present study examines the prevalence of ARB, ARGs and MGEs in SDBs, as well as WW and WWB, in both ICUs over a four-month period. The studied ICUs (NICU and AICU) are located in different buildings of the Grenoble Alpes University Hospital, allowing for the first time, the study and comparison of WW collecting waste from different hospital environments including 2 ICUs that apply different sink drain disinfection protocols and which host different types of patients (neonates versus adults). We applied a culturing approach that was focused on the isolation of CPEs and GRE, while characterization of the microbiota was achieved through complete sequencing of the 16S rRNA genes. The resistome was assessed through high-throughput qPCR targeting 86 ARGs and MGEs, with the objective of elucidating the dynamics of the SDB and WW microbiota and their resistome in response to disinfection measures at the NICU.

## 2. Material and methods

### 2.1. Location and setting

Grenoble Alpes University Hospital is the reference university hospital of the Isère department (South-East region of France). It is distributed across various sites and serves a population of 675,000 inhabitants with 2,074 hospital beds and a total of 431,893 hospitalization days in 2022. There are 26 beds in the NICU (including 10 beds on level 3) and 20 beds in the medical AICU localized in two different buildings and sites. Single or double rooms are present in the NICU while there are only single rooms in the AICU.

The sink drains of the sinks analyzed in our study were located in both the NICU and the AICU patients’ rooms. At both the NICU and the AICU, the patient surroundings and the room’s sink are cleaned daily with diluted (10 mL into 4 L of water) Surfanios Premium detergent/disinfectant (Surfanios Premium, Laboratoires ANIOS) and floors are cleaned daily using water and a microfiber gauze. At the NICU, sink drains are disinfected daily with 20 mL of undiluted Surfanios Premium and sink traps are renewed every six months. At the AICU, sink drains are only disinfected (with 20 mL of undiluted Surfanios Premium) during the stay of a patient that is colonized or infected with a CPE or GRE and at the exit of a patient with contact precaution (Lepelletier et al., 2015).

Screening of patients for CPEs and GRE was carried out according to the following protocol: at the NICU, CPE carriers and contact patients *i.e*., patients exposed to a CPE or GRE carrier, were screened by rectal swab. Upon admission to the AICU, patients with the following criteria are considered to be at risk of carrying CPE or GRE and are therefore screened by rectal swab: (i) patients with a history of abroad (foreign countries) hospitalization within the previous year; (ii) CPE or GRE carriers. Low risk patients are (i) patients that were hospitalized (> 24 h) at least once within the previous year; (ii) patients that were transferred from another hospital; (iii) patients that were transferred from nursing home, and (iv) contact patients *i.e*., patients exposed to a CPE or GRE carrier.

### 2.2. Collection and preparation of the samples

Biofilms and stagnant waters were sampled monthly from randomly selected sink drains of both wards (NICU and AICU): four SDBs were sampled from each ward at each sampling campaign. In addition, the same day of the sink drain sampling, hospital WW and hospital WWB were sampled from the outlets of both the NICU and the AICU. The WW pipes connected to the NICU also collect the WW from both the pediatric and the gynecologic units while the WW pipes connected to the AICU also collect the WW from surgical and neurosurgical ICUs, the emergency department, and the mortuary.

To collect the SDBs that formed along the sink drain walls, a Motin catheter (Peters Surgical) was introduced in the sink drain and used to detach the biofilm. Subsequently, the catheter was connected to a 50 mL syringe (Penta Ferte) to aspire the stagnant water containing the detached biofilm of the sink drain. The water was transferred to a PEHD sodium thiosulfate bottle (Gosseling) and transported to the laboratory for subsequent sample processing.

WW samples were collected by inserting an opened 50 mL centrifuge tube (Corning) attached to a string directly into the WW pipes, and processed on the same day. WWB samples were collected by inserting a closed 50 mL sterile centrifuge tube (Corning) attached to a string into the WW pipes for one month to let the biofilm develop on it. Consecutively, the tube was recovered, and the biofilm was detached by scratching it from the surface of the tube using a cell scraper the same day SDB and WW samples were collected. The biofilm was then dispersed in 1X Phosphate Buffered Saline (PBS: 137 mM NaCl, 2.7 mM KCl, 10 mM Na2HPO4, 1.8 mM KH2PO4, pH 7.4).

All samples were transferred into 50 mL tubes (VWR® Ultra High-Performance Centrifuge Tubes) and centrifuged at 7,000 revolutions per minute (rpm) for 15 minutes. The supernatants were discarded, and the pellets were dispersed in 5 mL of PBS. The resulting solutions were dispatched into three 2-mL Eppendorf tubes: two of the tubes were subsequently used for DNA extraction and molecular analyses while the content of the third one (1 mL of the original sample) was mixed with 1 mL of 40% glycerol, stored at -80°C, and subsequently used for the bacterial culture analyses.

### 2.3 Culture approach of the aerobic flora with focus on the isolation of CPE

#### 2.3.1. Isolation of bacterial colonies from the samples

Samples were cultured on non-selective chocolate agar plates supplemented with IsoVitalX (bioMérieux SA) to identify the total aerobic flora. Dilutions from 10^-1^ to 10^-7^ of each sample were prepared in 1X PBS, and 100 µL of the undiluted sample as well as 100 µL of each dilution were plated. The plates were incubated for 24 hours at 35°C under aerobic conditions, then for 24 hours at 22°C under aerobic conditions.

The samples were simultaneously cultured on selective media. To isolate *Pseudomonas aeruginosa*, 20 µL of undiluted sample were plated on Cetrimide agar plates (bioMérieux SA). To isolate carbapenem-resistant *P. aeruginosa* and *Enterobacterales*, 20 µL of undiluted sample were plated on ChromID CARBA agar (bioMérieux SA) and ChromID OXA-48 agar plates (bioMérieux SA), respectively. To isolate glycopeptide-resistant *Enterococci*, 20 µL of undiluted sample were plated on a ChromID VRE agar plate (bioMérieux SA). Each plate was incubated for 48 hours at 35°C under aerobic conditions. All the colonies presenting different morphologies, as assessed visually from the selective and non-selective media, were identified using matrix-assisted laser desorption/ionization time-of-flight mass spectrometry (MALDI-TOF MS).

#### 2.3.2 Identification of the isolated strains by MALDI-TOF MS

The colonies were picked from their respective agar plate using sterile pipette tips and spotted as thin films on their respective MALDI target positions (MSP 96 polished steel BC targets, Bruker Daltonics). Each bacterial film was overlaid with 1 µL of pre-portioned α-cyano- 4-hydroxycinnamic acid (HCCA) matrix (Bruker Daltonics), dried, and analyzed in a Microflex LT/SH Biotyper (Bruker Daltonics). The resulting spectra displaying the mass-to-charge values on the x-axis and the intensities on the y-axis were compared to a database containing 4,194 microorganism species (MBT IVD Library Resvison J, 2022, Brucker Daltonic), thus allowing the identification of the isolated strains based on their spectral matches to the reference profiles present in the database.

#### 2.3.2. Antibiotic susceptibility testing

Antibiograms were performed on the Enterobacterales and P. aeruginosa identified on the CARBA or OXA-48 plates, using the Kirby-Bauer disc diffusion method on Mueller-Hinton agar (MHA) plates (bioMérieux SA) and MHA-cloxacillin (bioMérieux SA). Briefly, bacterial suspensions were calibrated at 0.5 McFarland (around 1.5×108 colonies forming unit (CFU)/mL) and spread onto the surface of MHA and MHA cloxacillin plates. Discs impregnated with a known antibiotic at a precise concentration were placed on the agar surface. After 18-24 hours of incubation at 35°C, the growth inhibition diameters around the discs were measured to classify the bacteria as “susceptible”, “susceptible at increased exposure” or “resistant” to each tested antibiotic. Antibiotic susceptibility testing was performed according to the European guidelines (“eucast: AST of bacteria,” n.d.).

#### 2.3.4 Screening for CPE and carbapenemase producing *P. aeruginosa*

Screening tests were performed to detect CPE, further following the European guidelines (**Figure 1**). Bacteria that presented an inhibition diameter to ceftazidime-avibactam less than 10 millimeters, to temocillin less than 16 millimeters, or to meropenem or imipenem less than 22 millimeters were further tested to confirm their production of a carbapenemase. For *Serratia* spp., *Morganella* spp., and *Citrobacter freundii*, as wells as for *Enterobacter* spp. and *Providencia* spp., the inhibition diameter of meropenem or imipenem on MHA-cloxacillin plates must be less than 32 millimeters to eliminate the possibility of a resistance due to expression of the *ampC* gene that codes for a cephalosporinase.

**Figure 1:**
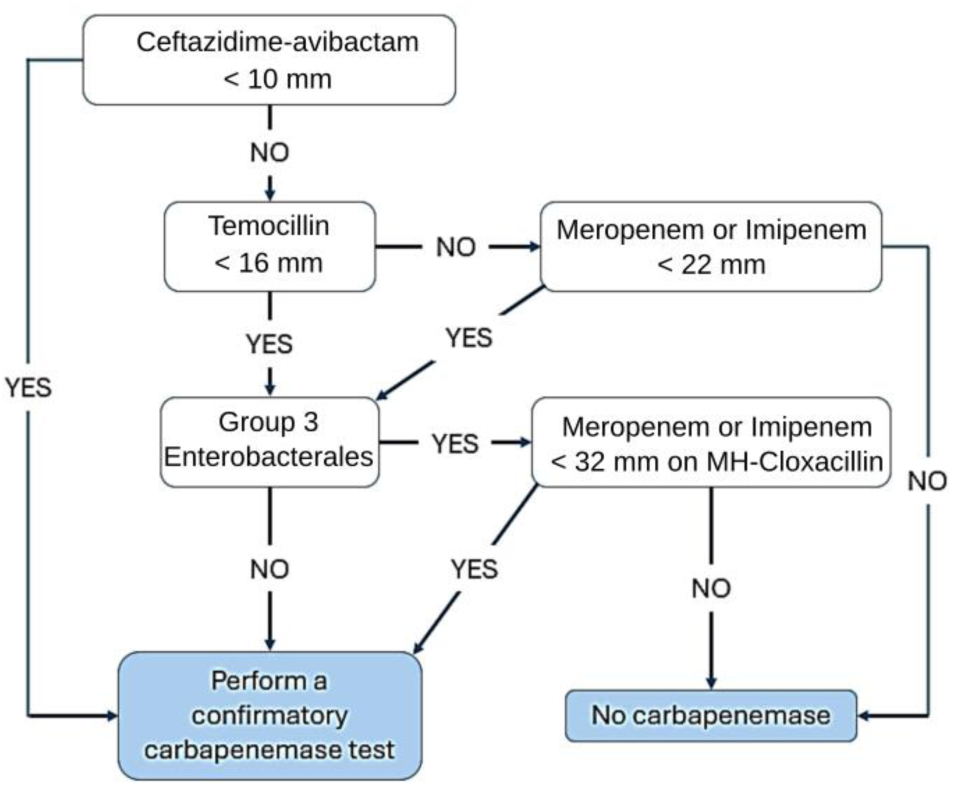
Schematization of the phenotypic screening of carbapenemase-producing *Enterobacterales* according to the 2023 recommendations from (“eucast: AST of bacteria,” n.d.).

Carbapenemase-producing *P. aeruginosa* strains were screened according to the French national guidelines (**Figure 2**). Strains with an inhibition area to imipenem less than 20 millimeters and less than 23 millimeters to ceftolozan-tazobactam were selected (“Algorithme_CARBA_PA_v2.pdf,” n.d.) For each selected strain, an immunochromatographic test (O.K.N.V.I resist-5 Coris test, Coris BioConcept) was carried out to detect the presence of the five major families of carbapenemases: OXA- 48, *Klebsiella pneumoniae* carbapenemase (KPC), New-Delhi metallo-β-lactamase (NDM), Verona integron–encoded metallo-β-lactamase (VIM) and imipenemase (IMP). If this test was negative, a confirmation test *i.e*., the colorimetric MAST CARBA PAcE assay (Mast Group) was performed to infirm or confirm a carbapenemase activity.

**Figure 2:**
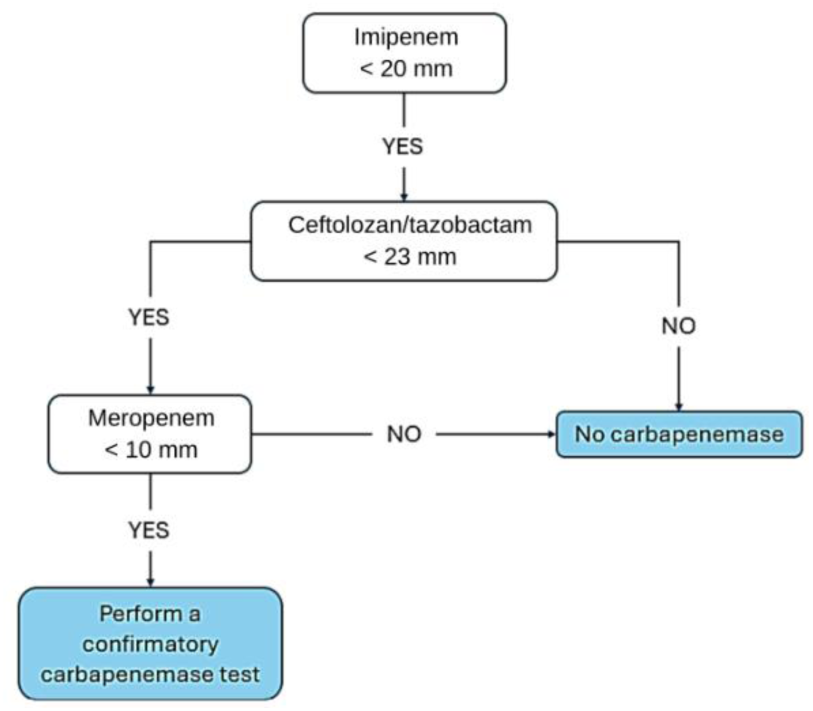
Schematization of the phenotypic screening of carbapenemase-producing *Pseudomonas aeruginosa*, according to the French « Centre National de Référence de la Résistance aux Antibiotiques » *(“Algorithme_CARBA_PA_v2.pdf,” n.d.)*.

### 2.4. Culture-independent approaches

#### 2.4.1. DNA extraction

Samples that had been stored at -80°C were thawed at room temperature for 10 min, centrifuged for 2 min at 20,800 g, room temperature, and the supernatant was discarded. The DNA extraction was performed using a FastDNA Spin kit for Soil (MP Biomedical’s products) according to the manufacturer’s instructions. Briefly, the pellets were transferred into 2 mL tubes containing lysing matrix E, a mixture of ceramic and silica particles designed to efficiently lyse soil organisms. Beat beating was performed for 5 min at the maximum speed (2,850 rpm) in a combination of lysing matrix E and sodium phosphate buffer pH 7.7, using a FastPrep instrument. The samples were then centrifuged for 15 min at 20, 800 g at room temperature. The DNA was purified from the supernatant with a silica-based GeneClean procedure using SPIN filters (MP Biomedicals). Concentrations of the eluted DNAs were determined by spectrophotometry using a NanoDrop 2000 apparatus (Thermo Scientific) and all the concentrations were adjusted to 50 ng/µL for subsequent quantitative PCR (qPCR) analysis.

#### 2.4.2. Microbiota analysis and full length 16S rRNA sequencing by PacBio

The study of the microbial diversity aims at revealing the species abundance and the taxonomic diversity in a microbial community based on the investigation of genetic materials. Taking advantage of long-read sequencing technologies, these materials can be investigated by single-molecule real-time (SMRT) sequencing on PacBio platforms generating circular consensus sequences (CCSs). CCS reads are filtered, clustered, and de-noised to generate full-length amplicon tags for species annotation and abundance analysis. Full-length 16S rRNA gene sequencing was performed by BMKGENE (Biomarker Technologies (BMK) GmbH). Construction of the library and sequencing were performed as described in the following. Total DNA was extracted from samples as mentioned previously. Specific primers with barcode were designed to target the full-length amplicon. PCR-amplified target sequences were purified, quantified, and homogenized to generate the SMRT bell library. Library quality control was performed on the libraries and the qualified ones were processed for sequencing on the PacBio Sequel II platform, which initially generated files in the bam format, then further converted in CCS files by smrtlink. The sequences were identified by barcode sequences, which resulted in a fastq sequences file for each sample. A total of 1,407,011 CCS reads were generated from 44 samples by barcode-based identification. A minimum of 2,416- and an average of 31,978 CCS reads were generated for each sample. One sample was excluded from the analysis due to low library quality.

#### 2.4.3 Bioinformatic analyses

Processing of the CCS files included the following three steps: (i) CCSs’ identification. CCS reads were identified based on barcode sequences by lima v1.7.0, generating raw- CCS files; (ii) CCSs’ filtration. Primer sequences were identified and removed by cutadapt 1.9.1. In the meantime, raw-CCSs were filtered based on their length (sequences between 1,200 and 1,650 bp were kept), which generated clean-CCSs; (iii) removal of chimeric sequences. Chimeric sequences were identified and removed by UCHIME v4.2, generating effective-CCSs. Bioinformatic analyses performed by BMKGENE included: (i) a feature identification OTUs/ASVs clustering; (ii) diversity analysis; (iii) differential analysis; (iv) correlation analysis and functional prediction.

### 2.5. High-throughput qPCR for the characterization of the resistome composition and normalized abundance

Nanoliter-scale quantitative PCRs were performed as described previously to quantify genes that confer resistance to antimicrobials (Buelow et al., 2023, 2020, 2018, 2017). We targeted 75 genes grouped in 16 resistance gene classes. These targeted genes included ARGs that are prevalent in the gut microbiota of healthy individuals (Hu et al., 2013), clinically relevant ARGs (extended-spectrum β-lactamases (ESBLs)-, carbapenemases-, and vancomycin resistance genes), heavy metals and quaternary ammonium compounds resistance genes suggested to favor cross and co–selection of ARGs in the environment (Langsrud et al., 2004). We also targeted 11 other genes including common transposase gene families, and class 1, 2 and 3 integron integrase genes, that are important vectors for ARGs in the clinics and which are often used as proxy for anthropogenic pollution (Gillings et al., 2015). Together, these 86 genes constituted what we named the resistome (Buelow et al., 2023, 2020, 2018, 2017). We also included primers targeting 16S rDNA. Primer design and validation prior to and after Biomark analysis was performed as described previously (Buelow et al., 2023, 2020, 2018, 2017). Real-time PCR analysis was performed using the 96.96 BioMark™ Dynamic Array for real-time PCR (Fluidigm Corporation, San Francisco, CA, U.S.A), as described previously. Thermal cycling and real-time imaging were performed at the INRAE Center of Research in Tours, France (UMR ISP1282, INRA Centre Val-de-Loire) and cycle threshold (Ct) values were calculated using the BioMark real-time PCR analysis software. Calculations for normalized and cumulative abundance of individual genes and allocated gene classes were performed as previously described (Buelow et al., 2023, 2020, 2018, 2017).

### 2.6. Analysis of the infection rate

Analysis of the infection rate at both the NICU and the AICU was performed between 01.01.2023 and 04.30.2023. All infants and adults hospitalized during this period were included in the analysis. Patients with a clinical sample and a microbiological identification of pathogenic bacteria were considered as infected. According to the French policy, patient consents were not required (retrospective study), and the data were declared to the Data Protection Officer of Grenoble Alpes University Hospital, France. The study protocol was reviewed by the clinical research direction of Grenoble Alpes University Hospital, France.

### 2.7. Statistical analyses

Normalized abundance of individual genes was accumulated per resistance gene class and Mann Whitney tests were performed with the scipy module v.1.13 to compare normalized cumulative abundance of the resistome between the NICU and the AICU SDBs, WW and WWB. A python script (v. 3.9) was realized to obtain the boxplots.

Logarithmic and Hellinger transformations were performed in R (v. 4.2) to correct for outliers in microbiota and resistome data. Redundancy analysis with 999 Monte Carlo permutations were then performed to test whether the microbiota explains the variations observed in the SDBs or the WW biofilm resistomes and to test for a significant correlation between the microbiome and the resistome in the respective samples. Principal coordinate analysis (PCA) was also performed in R and visualized with ggbiplot.

## 3. Results

The culturing approach was employed to characterize the total aerobic flora of SDBs collected at the NICU and the AICU from Grenoble Alpes University Hospital as well as samples collected from their respective WW outlets. Additionally, we screened for CPEs and GRE in these samples.

### 3.1. Culture-based analyses revealed diverse genera of aerobic bacteria in hospital sink drain biofilms

A total of 30 SDB samples were collected from both the NICU and the AICU. Over the four-month analysis period, a total of 18 SDB samples were collected at the NICU, including the sink drain of room #7, which was sampled twice. Twelve SDBs were collected from the AICU, including the sink drain of rooms 11 and 15, which were sampled twice, and the sink drain of rooms 12 and 14, which were sampled three times. Of the 18 SDB samples from the NICU, 72.2% (n = 13) were found to be contaminated with at least one bacterial species. Similarly, 58.3% (n = 7) of the 12 SDB samples from the AICU exhibited contamination. The bacterial cultures remained negative for the SDB samples from rooms 1, 12, 16, 22 and 26 of the NICU and for the SDB samples from rooms 12 (tested twice), 13, 14, and 15 of the AICU (**Figure 3A**). The aerobic flora cultivated from the SDBs using our culture approach exhibited low diversity, with an average of 3–6 (0–6) different bacterial species per SDB across the two ICUs. In both ICUs, the most frequently isolated genus was *Achromobacter* spp., identified in 72.2% (13/18) of the samples from the NICU and 50% (6/12) from the AICU (**Figure 3A**). Furthermore, *Achromobacter* spp. was the sole bacterial strain isolated from the SDBs of the NICU rooms 3 and 7 (**Figure 3A**). In addition to *Achromobacter* spp., other genera were also isolated from the SDBs of the NICU. *Serratia marcescens* was isolated from 44.4% of the samples (8/18), while *Pseudomonas aeruginosa* was isolated from 16.7% (3/18) of the samples. Additionally, *Ralstonia insidiosa*, *Stenotrophomonas maltophilia*, *Herbaspirillum huttiense*, and other species of *Pseudomonas* and *Serratia* genera were isolated at low frequencies (**Figure 3A**).

**Figure 3:**
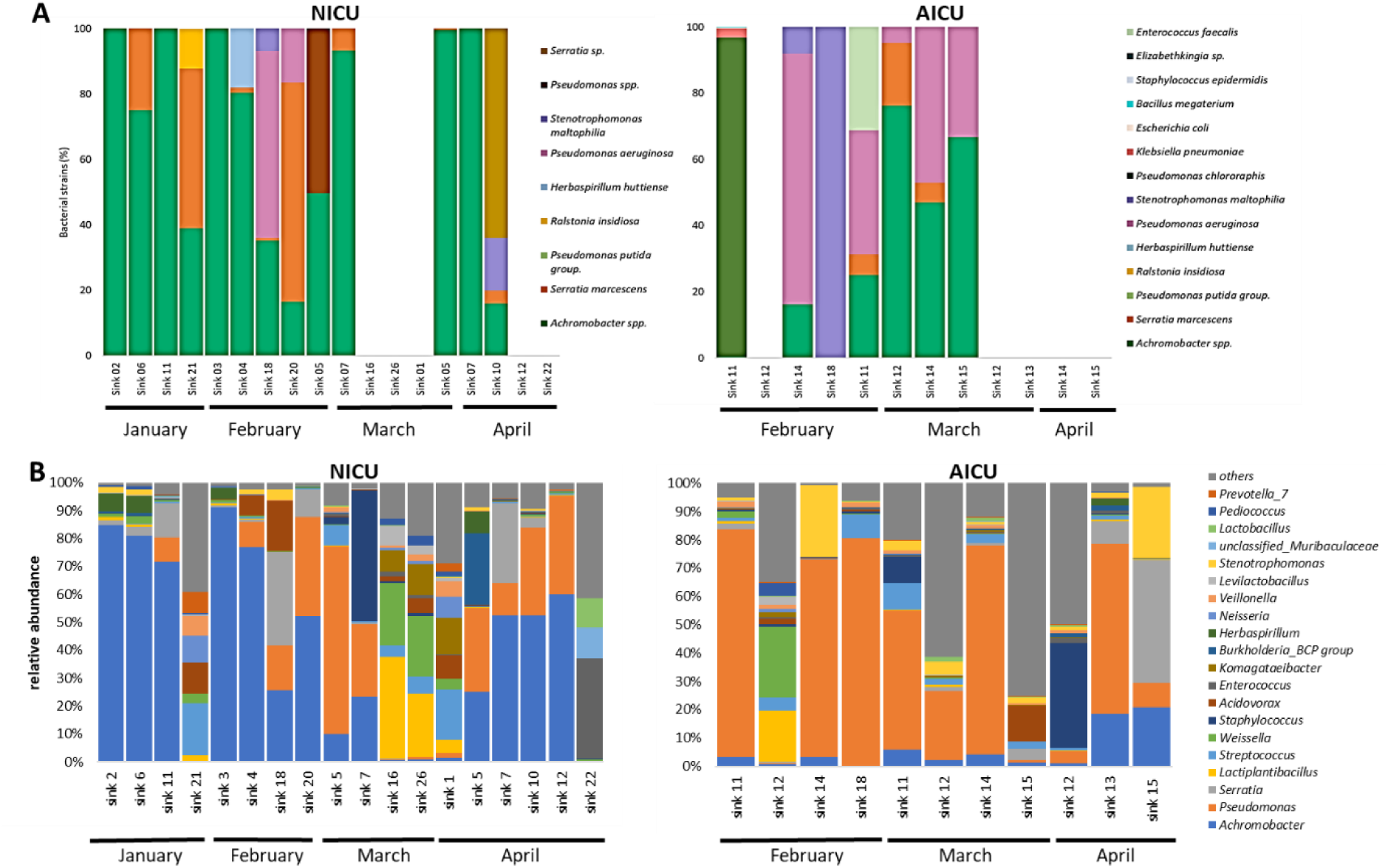
Comparison of the microbial species detected in sink drain biofilms of NICU *versus* AICU, as revealed by culture of the aerobic flora (A) and full-length 16S rRNA gene sequencing (B). A. The total aerobic flora was assessed by culture approaches of sink drain biofilms collected in the NICU *versus* the AICU over a 4-month period. Isolated bacterial strains were identified by MALDI-TOF-MS. CFUs/mL were determined and were expressed as percentages per sink drain biofilm. **B.** The microbiota of sink drain biofilms from the NICU *versus* the AICU was determined by full length 16S rRNA gene sequencing. The most abundant 20 genera are depicted while the remaining genera were grouped as “others”.

With the exception of *Achromobacter* spp., the SDBs of the AICU demonstrated a higher diversity of the total aerobic flora compared with that of the NICU, with the presence of *P. aeruginosa* (41.7% of the sinks; 5/12), *S. marcescens* (25%; 3/12), *Klebsiella pneumoniae*, *Escherichia coli*, *S. maltophilia*, *Staphylococcus epidermidis*, *Enterococcus faecalis*, and *Bacillus megaterium* at low frequencies. This study did not yield any isolations of CPEs nor GRE from the sink drain samples (**Figure 3A**).

### 3.2. The sink drain biofilm microbiota from the NICU differs from that of the AICU

The results from 16S rRNA gene sequencing (**Figure 3B**) were in strong agreement with those obtained from the cultures (**Figure 3A**), in particular for the NICU where the bacteria the most often encountered were *Achromobacter* spp. However, the sequencing approach revealed a greater diversity of bacterial genera in the biofilms of the sink drains than those detected by the culture approach. In general, the SDB microbiota of both the NICU and the AICU displayed a considerable degree of diversity between samples. Furthermore, for the same sink drains sampled on multiple occasions (NICU sink drain 7; AICU sink drains 1, 11, and 12), there was evidence of highly dynamic SDB communities. The microbiota of the NICU was found to differ from that of the AICU, with the former being dominated by the Gram-negative genera *Achromobacter*, *Serratia*, and *Acidovorax*, as well as the Gram-positive genera *Weisella*, *Lactiplantibacillus,* and *Levilactbacillus*. In contrast, the AICU microbiota was dominated by potential bacterial pathogens (Gram-positive and Gram-negative), including *Pseudomonas*, *Stenotrophomonas*, *Staphylococcus*, and *Klebsiella* (**Figure 3B** and **supplementary Figure 1**).

### 3.3 The sink drain biofilm resistome depends neither on the ICU nor on the disinfection protocol

No significant difference in normalized cumulative abundances was detected between the targeted gene class families for the analyzed SDBs from both ICUs and over time (**Figure 4**). Despite the dissimilarities in microbiota composition between the NICU and the AICU (**Figure 3**), no notable correlation was observed between the microbiota composition and the respective resistomes of the sink drain biofilms (**Supplementary Table 1**, **Supplementary** Figure 2, **Figure 4**). Furthermore, principal coordinate analysis confirmed that the SDBs of the two distinct ICUs, one caring for premature patients and the other for adults, exhibited a comparable resistome despite the disparate hygiene protocols in place (**Supplementary** Figure 2).

**Figure 4:**
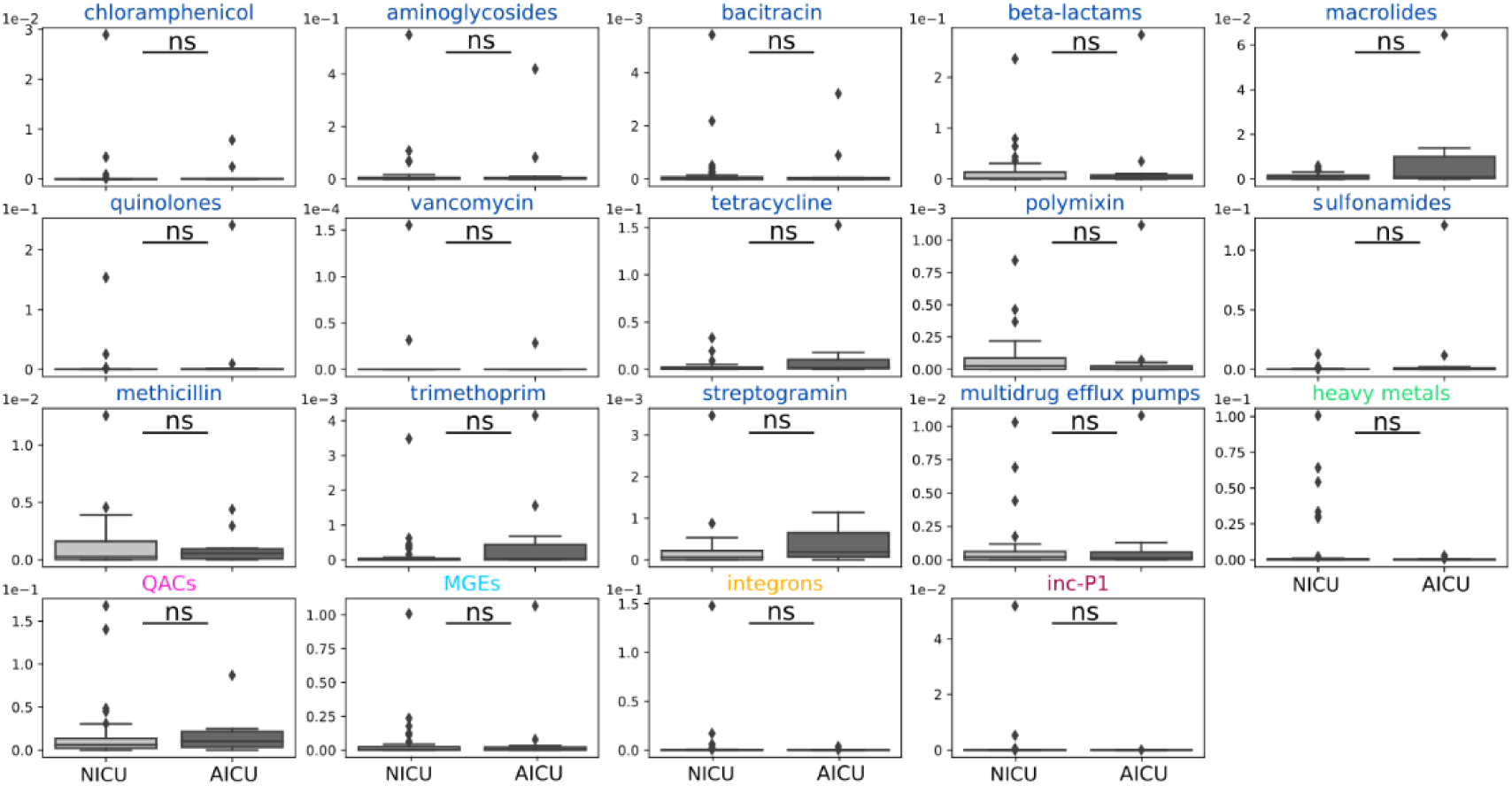
NICU *versus* AICU resistomes of sink drain biofilms. Individual genes were grouped in gene classes and cumulated normalized abundance per gene class was compared between the two ICUs using Mann Whitney tests to assess statistical differences. Dark blue: antibiotic resistance genes; green: heavy metals resistance genes; pink: resistance genes to quaternary ammonium compounds (QACs); light blue: transposase genes (mobile genetic elements; MGEs); yellow: integron integrase genes (class I, II and II); violet: IncP1 plasmid.

### 3.4. Culture-based analyses revealed the presence of a diverse range of aerobic bacteria in the WW of the respective ICUs

A total of four WW (which had not undergone any treatment prior to collection) and four WWB samples were collected at the NICU while four WW and only three WWB samples were collected at the AICU between January and April 2023. The bacterial strains isolated and cultured from the WW and WWB collected at both ICUs were more numerous and more diversified than those isolated from the SDBs. The average number of bacterial species per sample was 13 (range 10–16). It is noteworthy that *Aeromonas* spp. was isolated from all WW samples from the NICU as wells as from the AICU. The most prevalent bacterial species in the NICU WW and WWB samples were *E. coli*, *Klebsiella oxytoca*, and *Pseudomonas spp*. (including *P. aeruginosa*), while the most prevalent species at the AICU were *S. maltophilia*, *P. aeruginosa*, *S. marcescens*, and the *C. freundii* complex (**Figure 5A**).

**Figure 5:**
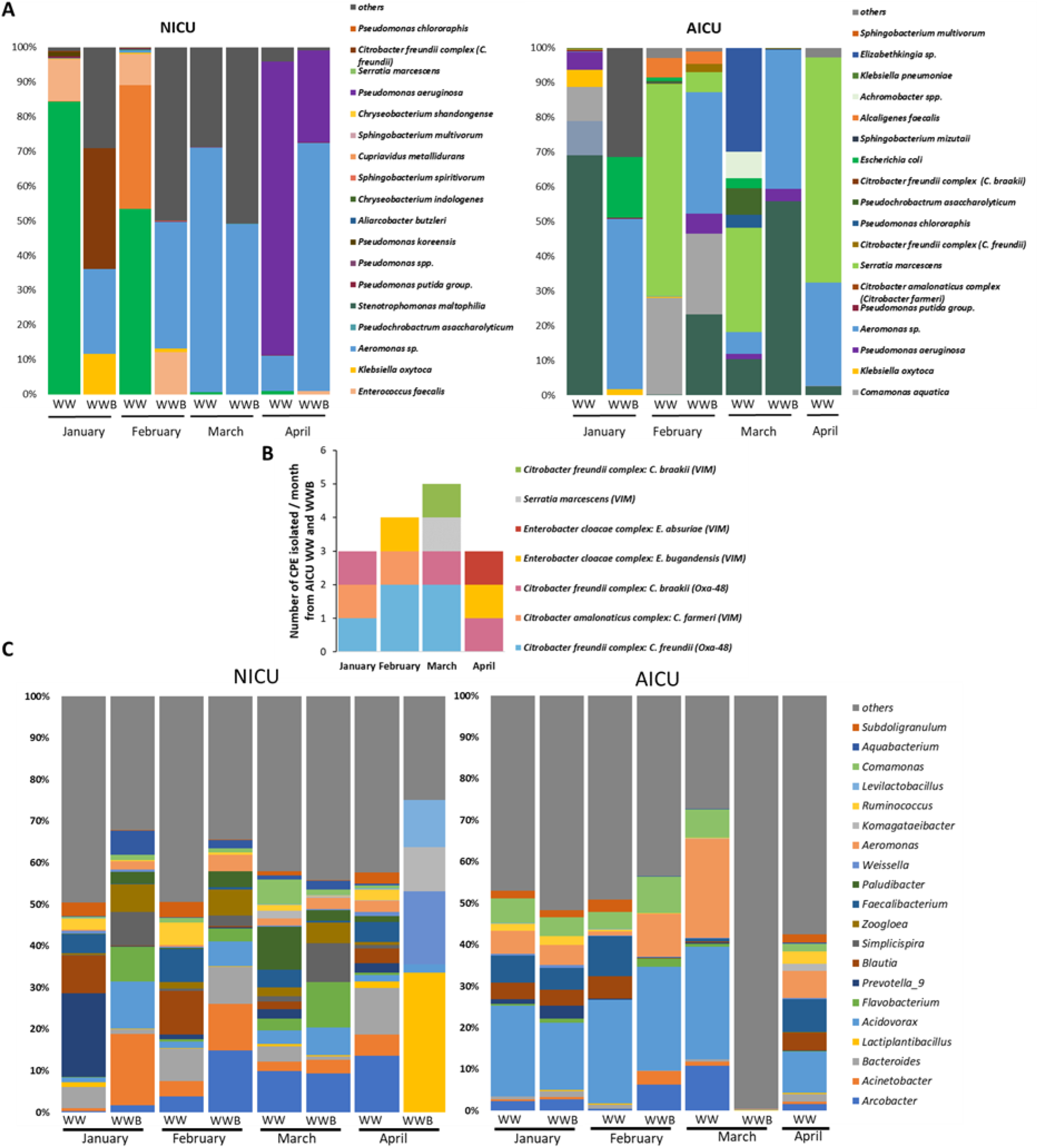
Analysis of waste water and waste water biofilm samples from both the NICU and the AICU. A. Total aerobic flora assessed by culture approaches of waste water and waste water sink drain biofilms from both the NICU and the AICU and sampled over a 4-month period (01-04.2023). Isolated bacterial strains were identified by MALDI-TOF-MS. CFUs/mL were determined and expressed as percentages per sink drain biofilm. The top 20 isolated species are depicted while the lesser abundant species are grouped and referred to as “others”. B. Carbapenemase producing *Enterobacterales* isolated from AICU WW and WWB by conventional culture. Total numbers of strains isolated per sampling time point (WW and WWB) and their respective identified carbapenemases. C. Microbiota of WW and WW biofilms at both the NICU and the AICU, as identified by full length 16S rRNA gene sequencing. The top 20 genera are depicted while the remaining genera are grouped and referred to as “others”.

### 3.5. Carbapenemase-producing *Enterobacterales* were mainly isolated from waste water and waste water biofilm samples from the AICU

We successfully isolated 25 strains of *P. aeruginosa* and *Enterobacterales* suspected of carbapenemase production on selective culture plates, which were subsequently tested for carbapenemase production. In total, one CPE strain (*C. freundii* complex-OXA- 48) and 15 CPE strains were isolated from either the WW or the WWBs collected from the WW drainpipes of the NICU and the AICU, respectively (**Figure 5B**). In the AICU, the majority of the CPE isolated strains (9/15) were belonging to the *Citrobacter freudii* complex. More precisely, we found: eight *Citrobacter freudii* complex carrying the OXA-48 carbapenemase, one *Citrobacter freudii* complex carrying the VIM carbapenemase. The remaining 6 CPE strains were two strains of the *Citrobacter amanilaticus* group carrying a VIM carbapenemase, three strains of the *Enterobacter cloacae* complex carrying VIM and one *Serratia marcescens* strain also carrying a VIM carbapenemase (**Figure 5B**). Together, these results demonstrated the presence of diverse CPEs in the WW collecting wastes from the AICU during the sample collection period.

### 3.6. The microbiota from both the NICU and the AICU, as revealed by 16S rRNA gene sequencing, were highly dynamic over the 4-month period of analysis

16S rRNA gene sequencing revealed that the microbiota of the NICU WW and WWB was more diverse than that of the AICU WW and WWB, and was characterized by genera such as *Arcobacter*, *Acinetobacter*, *Bacteroidetes*, *Flavobacterium*, and *Zoogleoa*. In contrast, the AICU WW and WWB microbiota were characterized by *Pseudomonas*, *Achromobacter*, *Acidovorax*, *Strenophomonas,* and *Aeromonas* (**Figure 5C** and **Supplementary** Figure 4). While the NICU and AICU WW and WWB microbiota signatures were more distinctly separated than those observed in the respective sink drain microbiota (**Figure 3B** and **Supplementary** Figure 1), both NICU and AICU WW and WWB microbiota signatures demonstrated high dynamism between the different sampling time points (**Figure 5C** and **Supplementary** Figure 4). The microbiota signatures of WW and WWBs were thus found to be in agreement with the culture-based determined aerobic flora, while also revealing a large reservoir of anaerobic Gram-negative and Gram-positive bacteria that were not isolated nor detected by the culture approach.

### 3.7. The WW and WW biofilm resistome of the AICU contains more resistance genes than that of the NICU

Accumulated normalized abundances of genes conferring resistance to quinolones and integron integrase genes were significantly higher in the AICU WW and WWB, compared to those from the NICU WW and WWB, while accumulated normalized abundance of genes conferring resistance to streptogramin, an antibiotic active against Gram-positive bacteria such as GRE or vancomycin-resistant *Staphylococcus aureus* (Reissier and Cattoir, 2021), was higher in the NICU WW and WWB (**Figure 6**). There was also a trend of higher accumulated normalized abundances of resistance genes to bacitracin, tetracyclins, sulfonamides and methicillin in the NICU WW and WWB, while there was a trend of higher accumulated normalized abundances of resistance genes to chloramphenicol, aminoglycosides, β- lactams, vancomycin, trimethoprim, quaternary ammonium compounds, as well as MGEs and incP1 plasmid in the AICU WW and WWB (**Figure 6**). Importantly, no significant correlation was found between the microbiota composition (**Figure 5C**) and the resistome in the NICU and AICU WW and WWB samples (**Figure 6**). Moreover, redundancy analysis (RDA) revealed no clear constraints that could explain the observed variance in the resistome dataset (**Supplementary Table 1**).

**Figure 6:**
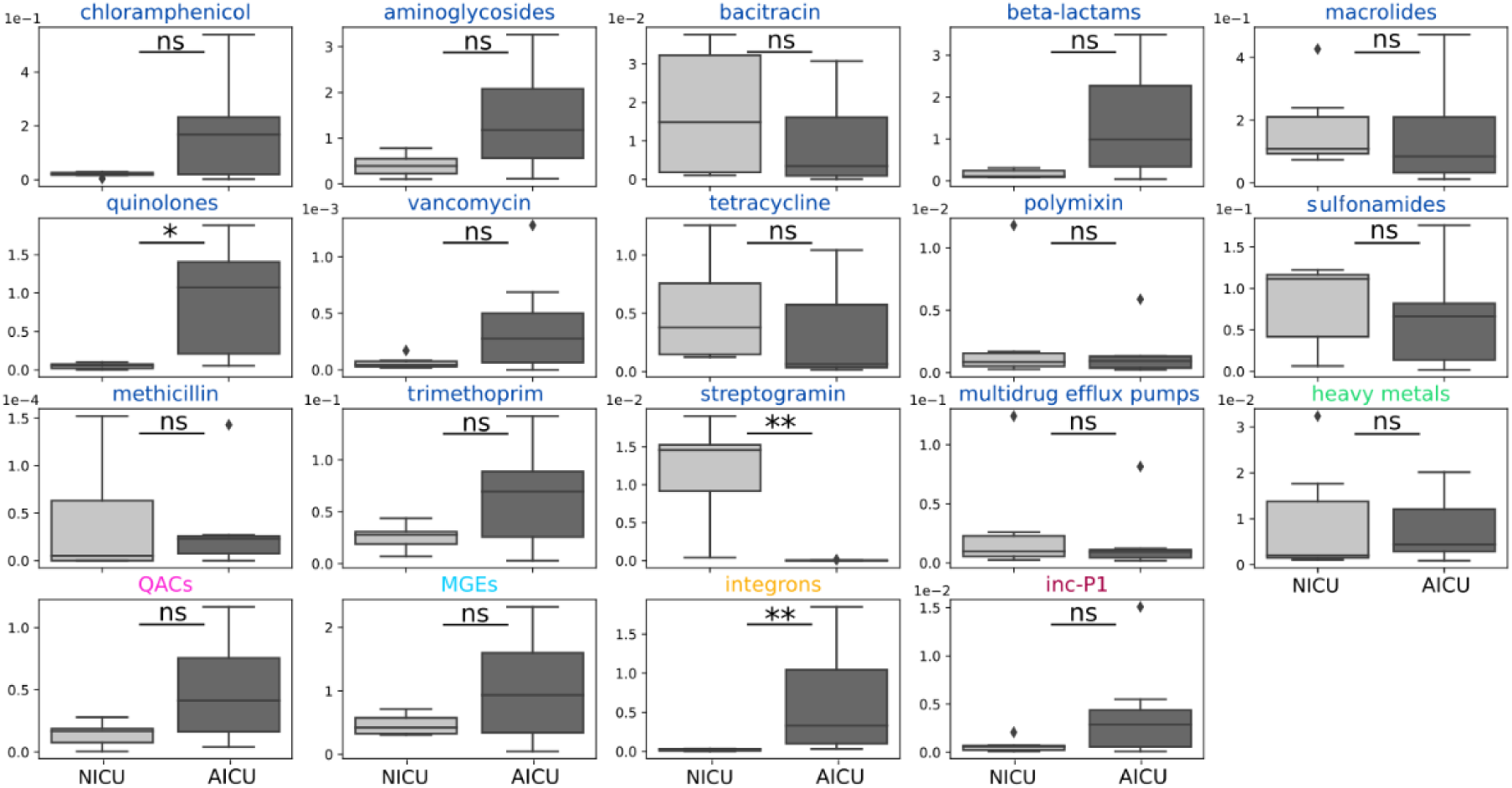
NICU *versus* AICU resistomes from WW and WW biofilms. Individual genes were grouped into gene classes and accumulated normalized abundance per gene class was compared between the two ICUs. To assess statistical differences, Mann Whitney tests were performed. The color code is the following: blue color, class of antibiotic the targeted genes confer resistance to; green, heavy metals resistance genes; pink, resistance genes to quaternary ammonium compounds (QACs); light blue, transposase genes (mobile genetic elements; MGEs); yellow, integron integrase genes (class I, II and II); violet, IncP1 plasmid. (*, *p*<0.05; **, *p*<0.01).

### 3.8. Infection incidences in hospitalized patients in both ICUs during the period of study

The number of hospitalized patients in each ICU during the period of study as well as the infection incidences were also recorded (**Table 1**). At the AICU, 29.2 % of hospitalized patients were infected with pathogenic bacteria *versus* 8.6 % at the NICU. At the AICU, 28 patients were infected by *Enterobacterales versus* 5 neonates at the NICU between January and April 2023. No CPE were isolated from the patients that were screened during the respective time period in both wards. Infections in patients at the AICU were due to *Enterobacterales* (28 out of 71 patients), *Enterococcus* spp. (11 infected patients) or *Pseudomonas* spp. (11 infected patients), while infections in neonates were due to *Enterobacterales* in five neonates, six neonates were infected with *Enterococcus spp*., and none with *Pseudomonas spp*. (**Table 1**). While the strains isolated from patients *versus* SDBs during our study could not be compared for either ICU, the higher abundance of infections in patients due to *Enterobacterales*, *Enterococcus* spp. or *Pseudomonas* spp. from the AICU SDBs compared to the NICU SDBs is notable and corresponds to the overall higher rate of isolation of these pathogens in AICU patients during our study period (**Table 1**).

**Table 1:** Infection incidences in patients hospitalized in the NICU *versus* the AICU during the period of our study. IQR= interquartile.

| Category | NICU (n = 243 patients) | AICU (n = 243 patients) |
| --- | --- | --- |
| **Gender distribution** | 131 boys (53.91%),<br>112 girls (46.09%) | 163 men (67.08%),<br>80 women (32.92%) |
| **Median age** | N/A | 62.77 years (IQR: 50.44–71.69) |
| **Median length of stay** | 3.16 days (IQR: 1.23–10.50) | 4.04 days (IQR: 1.98–8.70) |
| **Number of infected patients:** | 21 (8.64%) | 71 (29.22%) |
| *Enterobacterales* | 5 (2.06%) | 28 (11.52%) |
| *Enterococcus* spp. | 6 (2.47%) | 11 (4.57%) |
| *Pseudomonas* spp. | None | 11 (4.57%) |
| GRE or CPE | None | None |

## 4. Discussion

The objective of this study was to examine the microbiota composition and the resistome of SDBs and WW systems in both the NICU and the AICU of the Grenoble Alpes University Hospital. The findings suggest a significant divergence in microbial flora between the NICU and the AICU settings, and also indicate an absence of correlation between the microbiota and the resistome profiles.

A culture-based analysis revealed that the SDBs in the NICU and the AICU harbored diverse genera of aerobic bacteria, with *Achromobacter* spp. being the most frequently isolated in both settings. It is notable that the NICU exhibited a lower diversity of bacterial species compared to the AICU, which demonstrated a greater prevalence of potential pathogens, including *Pseudomonas* and *Klebsiella*. These findings are consistent with those of previous studies which have indicated that intensive care environments can harbor significant microbial diversity, often dominated by antibiotic-resistant species (Aracil-Gisbert et al., 2024; Constantinides et al., 2020, 2020). However, while the SDBs from the AICU demonstrated a more varied microbiota, no CPEs were isolated from either the NICU or the AICU SDBs. Isolation of CPEs from environmental samples such as SDBs is challenging and our culture approach might have missed the presence of CPEs that may have a low MIC for carbapenems, or may carry carbapenem resistance genes on MGEs that were not expressed during our culture conditions (Dierikx et al., 2022). It is noteworthy that in the AICU, a total of 29.2% of the adults tested positive for infections caused by *Enterobacterales*, *Enterococcus* spp., and *Pseudomonas* spp. (**Table 1**). Notably, these pathogens were also isolated by our culture approach from the SDBs at the AICU. The presence of Gram-negative bacteria, particularly *S. marcescens* and *P. aeruginosa*, in the SDBs underscores two key concerns: firstly, the potential for transmission to vulnerable patients within healthcare facilities, and secondly, the importance of addressing this issue in infection control practices (Lalancette et al., 2017; Millán-Lou et al., 2021). Additional research is required to ascertain whether the strains isolated from the sinks are indeed the strains responsible for patient infections. For example, a recent study employing whole genome sequencing demonstrated that *P. aeruginosa* strains isolated from patients differed from those that colonized the sink drains in a hospital ICU (Couchoud et al., 2023). Overall, our findings indicate that the aerobic SDB microbiota identified by cultures seems indicative of the bacterial pathogens that are likely to infect and/or be carried by patients in the respective ICUs. Whether these strains are transmitted from patients to the SDBs or *vice versa* or not at all, will require further studies.

Our analysis of the resistome showed that there were no statistically significant differences in the abundance of resistance genes between the NICU and the AICU SDBs over time. This finding questions our initial hypothesis that disinfection protocols would result in a discernible impact on the resistome.

Biofilms provide protection to bacterial cells, rendering them resistant to the hostile environment created by detergent/disinfectant exposure. Our observations are consistent with previous studies that have reported the resilience of multispecies biofilm communities to various biocides, including chlorhexidine, benzalkonium chloride, peracetic acid, and chlorine (Kara et al., 2006; Schwering et al., 2013; van der Veen and Abee, 2011). The persistence of these biofilms despite the use of disinfectants, highlights the need for alternative strategies and approaches to effectively target and control biofilm-associated pathogens in healthcare settings (Karygianni et al., 2020, 2020; Maillard and Centeleghe, 2023; Volling et al., 2021). The lack of difference in the normalized abundance of the resistome despite notable differences in the microbiota may be explained by the fact that the analyzed DNA for both resistome and microbiome analyses is not limited to that of live and intact bacterial cells, but also includes extracellular DNA (eDNA) from the biofilm matrix. eDNA has been identified as a source for ARGs and MGEs in biofilms (Hannan et al., 2010; Murray et al., 2022; Sivalingam et al., 2020). Moreover, it is not uncommon for ARGs to be carried by broad-host range MGEs, such as IncP1 and IncPromA plasmids, which can be harbored by both Gram-negative and Gram-positive bacteria (Klümper et al., 2015). The persistence of eDNA in our analyzed SDBs and MGEs associated with ARGs may explain why their normalized abundance remains unchanged despite differences in the relative abundance of bacterial genera. In light of these findings, it can be proposed that the mere presence of specific bacterial species in these environments does not necessarily dictate the abundance of resistance genes. Our findings additionally indicate that ARGs may persist in the environment independently of the bacteria that carry them, which could complicate the management of antibiotic resistance in clinical settings (Catho et al., 2024; Ramos-Castaneda et al., 2020). The relationship between microbiota and resistome is often intricate and susceptible to be influenced by a multitude of environmental factors, which may not be adequately represented in the available dataset. For instance, factors such as sink drain usage, antibiotic usage, patient characteristics, or hospital conditions may contribute to variations in the resistome, potentially obscuring direct correlations with microbiota composition.

A greater diversity of bacterial species was observed in the WW samples than in the SDBs, with a notable prevalence of CPEs, particularly at the AICU. The dynamic nature of the WW microbiota highlights the necessity for monitoring these environments, as they may serve as crucial reservoirs for ARB that can disseminate within hospital settings as well as the environment (Buelow et al., 2023; Dias et al., 2020; Snell et al., 2024). So far, monitoring and analysis of WW collecting wastes from different patient populations such as neonates *versus* adults with different health care protocols (including antibiotic and medical prescription practices), and which correspond to different sink drain disinfection protocols, has not been performed yet. The differences detected in the aerobic and total microbiota, the overall resistome composition, and normalized abundances of resistance genes between the NICU and the ICU WW, as well as the presence or not of CPEs are likely to represent the very different clinical environments including the types of patients, and associated health care and hygiene protocols. Finally, identification of carbapenemase-producing strains in both WW and WWBs highlights the persistent challenge posed by ARB in hospital WW systems. Nevertheless, the question of whether WW can be used as an indicator for patient colonization with CPEs remains unanswered. Further investigation is required to determine whether the presence of these strains in WW indicates actual patient colonization or a risk of infection for patients with CPEs. In our study, at the NICU, patients were rarely screened for CPEs, and at the AICU this screening was also not performed systemically, but only for patients at risk, hence CPEs that were carried by patients during our study period might not have been detected. To address this question, a prospective study comparing different health care services should be performed to systematically screen all patients admitted to the NICU and the AICU for CPE and GRE and to perform a continuous SDB and WW screening using the ensemble of culture-dependent and culture-independent methods.

Together, our results indicate that, while disinfection protocols in the NICU may constrain the diversity of the SDB microbiota, they do not markedly influence the associated resistome. This prompts the need to re-examine the efficacy of current disinfection protocols and to consider the potential benefits of a more sophisticated approach to infection control that takes into account the complexities of microbial communities and their resistance profiles. Future studies should explore the potential for targeted disinfection strategies that can effectively reduce both the microbial diversity and the antibiotic resistance (the” resistome”) in critical care environments, and further focus on WW as suitable surveillance tool for the prediction of bacterial pathogens’ presence in the respective hospital environments.

## 5. Conclusions

- The microbiota composition of the NICU and the AICU SDBs were found to be distinct, while their resistome composition was similar.
- Disinfection does not affect the resistome at the NICU. Regular disinfection of NICU sink drains with surfactants did not significantly impact the overall resistome of SDBs compared to the resistome of SDBs from the AICU, which was not exposed to routine disinfection.
- The WW microbiota was very dynamic in both ICUs over time but significantly different between the NICI and the AICU.
- The AICU WW samples exhibited a markedly higher abundance of genes conferring resistance to quinolones, and integron integrase genes, whereas the NICU WW samples demonstrated elevated levels of genes resistant to streptogramin. These observations indicate notable distinctions in the composition of the respective ICU WW resistomes.
- No carbapenemase-producing *Enterobacterales* (CPEs) were isolated from SDBs. In contrast, 15 CPE strains were identified in AICU WW, and one CPE strain was isolated from NICU WW, underscoring the necessity of WW monitoring for ARB.
- The present study provides evidence that sink drain biofilms are resilient to disinfection interventions with quaternary ammonium compounds at the University Hospital Grenoble Alpes.

## Data Availability

All data produced in the present study are available upon reasonable request to the authors
Sequencing data are available online at the ENA database, accession number: PRJEB83107

## Acknowledgements

We would like to acknowledge (i) the hospital staff and nurses from the hygiene department, Nadia Kurtz, Gwenaellea Collard and Jennifer Charbonnier, for their assistance during collection of the samples; (ii) the technical team of the hospital Michel Gomez and Nicolas Blanc; (iii) the direction of the TIMC laboratory for supporting this research financially with the TIMC Emergence grant and the J.M.-R. student’s gratification; and (iv) the Asposan executive board for the financial support of this study.

Authors contributions: **E.B.** and **C.L.** designed the study; **A.H.** and **J.M.-R.** performed the experiments, analyzed the data and wrote the manuscript’s first draft; **M.R.M**. performed the statistical and bioinformatical analyses, and designed the figures; **C.T.M.** and **C.L.** allowed access to samples and the patients and hospital environment’s information; **A.H.** and **M.M**. allowed access and guidance for bacterial cultures and analyses; **A.H., J.M.-R., P.M., M.M., C.M., C.L.,** and **E.B**. reviewed the data and edited the manuscript. **C.M., C.L., E.B.,** and **A.H.** supervised the student.

## Data availability

Raw and assembled 16S rRNA sequences are available under accession number PRJEB83107.

## Notes

### Competing Interest Statement

The authors have declared no competing interest.

### Funding Statement

The study was funded by the TIMC laboratory and by the Association-Asposan, Montbonnot-Saint-Martin, France

### Author Declarations

According to the French law, submission to an IRB and patient consents are not required for retrospective studies based on existing healthcare systems data. However, in accordance with the Privacy of Individually Identifiable Health Information law, the research was declared to the Data Protection Officer of Grenoble Alpes University Hospital, France. The study protocol was reviewed by the clinical research direction of Grenoble Alpes University Hospital, France. The individual-level data had been de-identified (hence anonymized) prior to use in our study.

## References

1. Abe, K., Nomura, N., Suzuki, S., 2020. Biofilms: hot spots of horizontal gene transfer (HGT) in aquatic environments, with a focus on a new HGT mechanism. FEMS Microbiology Ecology 96. 10.1093/femsec/fiaa031

2. Algorithme_CARBA_PA_v2.pdf, n.d.

3. Aracil-Gisbert, S., Fernández-De-Bobadilla, M.D., Guerra-Pinto, N., Serrano-Calleja, S., Pérez-Cobas, A.E., Soriano, C., de Pablo, R., Lanza, V.F., Pérez-Viso, B., Reuters, S., Hasman, H., Cantón, R., Baquero, F., Coque, T.M., 2024. The ICU environment contributes to the endemicity of the “*Serratia marcescens* complex” in the hospital setting. mBio 15, e03054–23. 10.1128/mbio.03054-23

4. Balcazar, J.L., Subirats, J., Borrego, C.M., 2015. The role of biofilms as environmental reservoirs of antibiotic resistance. Front.Microbiol. 6, 1216. 10.3389/fmicb.2015.01216

5. Bourdin, T., Benoit, M.-È., Prévost, M., Charron, D., Quach, C., Déziel, E., Constant, P., Bédard, E., 2024. Disinfection of sink drains to reduce a source of three opportunistic pathogens, during *Serratia marcescens* clusters in a neonatal intensive care unit. PLoS One 19, e0304378. 10.1371/journal.pone.0304378

6. Buelow, E., Bayjanov, J.R., Majoor, E., Willems, R.J.L., Bonten, M.J.M., Schmitt, H., van Schaik, W., 2018. Limited influence of hospital wastewater on the microbiome and resistome of wastewater in a community sewerage system. FEMS Microbiol.Ecol. 10.1093/femsec/fiy087

7. Buelow, E., Bello Gonzalez, T.D.J., Fuentes, S., de Steenhuijsen Piters, W.A.A., Lahti, L., Bayjanov, J.R., Majoor, E.A.M., Braat, J.C., van Mourik, M.S.M., Oostdijk, E.A.N., Willems, R.J.L., Bonten, M.J.M., van Passel, M.W.J., Smidt, H., van Schaik, W., 2017. Comparative gut microbiota and resistome profiling of intensive care patients receiving selective digestive tract decontamination and healthy subjects. Microbiome 5, 88–017-0309-z. 10.1186/s40168-017-0309-z

8. Buelow, E., Dauga, C., Carrion, C., Mathé-Hubert, H., Achaibou, S., Gaschet, M., Jové, T., Chesneau, O., Kennedy, S.P., Ploy, M.-C., Da Re, S., Dagot, C., 2023. Hospital and urban wastewaters shape the matrix and active resistome of environmental biofilms. Water Research 244, 120408. 10.1016/j.watres.2023.120408

9. Buelow, E., Rico, A., Gaschet, M., Lourenço, J., Kennedy, S.P., Wiest, L., Ploy, M.-C., Dagot, C., 2020. Hospital discharges in urban sanitation systems: Long-term monitoring of wastewater resistome and microbiota in relationship to their eco-exposome. Water Research X 7, 100045. 10.1016/j.wroa.2020.100045

10. Catho, G., Cave, C., Grant, R., Carry, J., Martin, Y., Renzi, G., Nguyen, A., Buetti, N., Schrenzel, J., Harbarth, S., 2024. Controlling the hospital aquatic reservoir of multidrug-resistant organisms: a cross-sectional study followed by a nested randomized trial of sink decontamination. Clinical Microbiology and Infection 30, 1049–1054. 10.1016/j.cmi.2024.05.008

11. Constantinides, B., Chau, K.K., Quan, T.P., Rodger, G., Andersson, M.I., Jeffery, K., Lipworth, S., Gweon, H.S., Peniket, A., Pike, G., Millo, J., Byukusenge, M., Holdaway, M., Gibbons, C., Mathers, A.J., Crook, D.W., Peto, T.E.A., Walker, A.S., Stoesser, N., 2020. Genomic surveillance of *Escherichia coli* and *Klebsiella spp.* in hospital sink drains and patients. Microb Genom 6, mgen000391. 10.1099/mgen.0.000391

12. Couchoud, C., Bertrand, X., Bourgeon, M., Piton, G., Valot, B., Hocquet, D., 2023. Genome-based typing reveals rare events of patient contamination with *Pseudomonas aeruginosa* from other patients and sink traps in a medical intensive care unit. Journal of Hospital Infection 134, 63–70. 10.1016/j.jhin.2023.01.010

13. De Geyter, D., Vanstokstraeten, R., Crombé, F., Tommassen, J., Wybo, I., Piérard, D., 2021. Sink drains as reservoirs of VIM-2 metallo-β-lactamase-producing *Pseudomonas aeruginosa* in a Belgian intensive care unit: relation to patients investigated by whole-genome sequencing. Journal of Hospital Infection 115, 75–82. 10.1016/j.jhin.2021.05.010

14. de Jonge, E., de Boer, M.G.J., van Essen, E.H.R., Dogterom-Ballering, H.C.M., Veldkamp, K.E., 2019. Effects of a disinfection device on colonization of sink drains and patients during a prolonged outbreak of multidrug-resistant *Pseudomonas aeruginosa* in an intensive care unit. J Hosp Infect 102, 70–74. 10.1016/j.jhin.2019.01.003

15. Dias, M.F., da Rocha Fernandes, G., Cristina de Paiva, M., Christina de Matos Salim, A., Santos, A.B., Amaral Nascimento, A.M., 2020. Exploring the resistome, virulome and microbiome of drinking water in environmental and clinical settings. Water Research 174, 115630. 10.1016/j.watres.2020.115630

16. Dierikx, C., Börjesson, S., Perrin-Guyomard, A., Haenni, M., Norström, M., Divon, H.H., Ilag, H.K., Granier, S.A., Hammerum, A., Kjeldgaard, J.S., Pauly, N., Randall, L., Anjum, M.F., Smialowska, A., Franco, A., Veldman, K., Slettemeås, J.S., 2022. A European multicenter evaluation study to investigate the performance on commercially available selective agar plates for the detection of carbapenemase producing *Enterobacteriaceae*. Journal of Microbiological Methods 193, 106418. 10.1016/j.mimet.2022.106418

17. eucast: AST of bacteria [WWW Document], n.d. URL https://www.eucast.org/ast_of_bacteria (accessed 11.19.24).

18. Gillings, M.R., Gaze, W.H., Pruden, A., Smalla, K., Tiedje, J.M., Zhu, Y.G., 2015. Using the class 1 integron-integrase gene as a proxy for anthropogenic pollution. ISME J. 9, 1269–1279. 10.1038/ismej.2014.226

19. Hannan, S., Ready, D., Jasni, A.S., Rogers, M., Pratten, J., Roberts, A.P., 2010. Transfer of antibiotic resistance by transformation with eDNA within oral biofilms. FEMS Immunology & Medical Microbiology 59, 345–349. 10.1111/j.1574-695X.2010.00661.x

20. Hopman, J., Tostmann, A., Wertheim, H., Bos, M., Kolwijck, E., Akkermans, R., Sturm, P., Voss, A., Pickkers, P., vd Hoeven, H., 2017. Reduced rate of intensive care unit acquired gram-negative bacilli after removal of sinks and introduction of ‘water-free’ patient care. Antimicrob Resist Infect Control 6, 59. 10.1186/s13756-017-0213-0

21. Hu, Y., Yang, X., Qin, J., Lu, N., Cheng, G., Wu, N., Pan, Y., Li, J., Zhu, L., Wang, X., Meng, Z., Zhao, F., Liu, D., Ma, J., Qin, N., Xiang, C., Xiao, Y., Li, L., Yang, H., Wang, J., Yang, R., Gao, G.F., Wang, J., Zhu, B., 2013. Metagenome-wide analysis of antibiotic resistance genes in a large cohort of human gut microbiota. Nat.Commun. 4, 2151. 10.1038/ncomms3151;

22. Kara, D., Luppens, S.B.I., ten Cate, J.M., 2006. Differences between single- and dual-species biofilms of *Streptococcus mutans* and *Veillonella parvula* in growth, acidogenicity and susceptibility to chlorhexidine. European Journal of Oral Sciences 114, 58–63. 10.1111/j.1600-0722.2006.00262.x

23. Karygianni, L., Ren, Z., Koo, H., Thurnheer, T., 2020. Biofilm Matrixome: Extracellular Components in Structured Microbial Communities. Trends in Microbiology 28, 668–681. 10.1016/j.tim.2020.03.016

24. Klümper, U., Riber, L., Dechesne, A., Sannazzarro, A., Hansen, L.H., Sørensen, S.J., Smets, B.F., 2015. Broad host range plasmids can invade an unexpectedly diverse fraction of a soil bacterial community. The ISME Journal 9, 934–945. 10.1038/ismej.2014.191

25. Lalancette, C., Charron, D., Laferrière, C., Dolcé, P., Déziel, E., Prévost, M., Bédard, E., 2017. Hospital Drains as Reservoirs of *Pseudomonas aeruginosa*: Multiple-Locus Variable-Number of Tandem Repeats Analysis Genotypes Recovered from Faucets, Sink Surfaces and Patients. Pathogens 6, 36. 10.3390/pathogens6030036

26. Langsrud, S., Sundheim, G., Holck, A.L., 2004. Cross-resistance to antibiotics of *Escherichia coli* adapted to benzalkonium chloride or exposed to stress-inducers. J.Appl.Microbiol. 96, 201–208.

27. Ledwoch, K., Robertson, A., Lauran, J., Norville, P., Maillard, J.-Y., 2020. It’s a trap! The development of a versatile drain biofilm model and its susceptibility to disinfection. J Hosp Infect 106, 757– 764. 10.1016/j.jhin.2020.08.010

28. Lepelletier, D., Berthelot, P., Lucet, J.-C., Fournier, S., Jarlier, V., Grandbastien, B., National Working Group, 2015. French recommendations for the prevention of “emerging extensively drug- resistant bacteria” (eXDR) cross-transmission. J Hosp Infect 90, 186–195. 10.1016/j.jhin.2015.04.002

29. Low, J.M., Chan, M., Low, J.L., Chua, M.C.W., Lee, J.H., 2024. The impact of sink removal and other water-free interventions in intensive care units on water-borne healthcare-associated infections: a systematic review. Journal of Hospital Infection 150, 61–71. 10.1016/j.jhin.2024.05.012

30. Maillard, J.-Y., Centeleghe, I., 2023. How biofilm changes our understanding of cleaning and disinfection. Antimicrobial Resistance & Infection Control 12, 95. 10.1186/s13756-023-01290-4

31. Millán-Lou, M.I., López, C., Bueno, J., Pérez-Laguna, V., Lapresta, C., Fuertes, M.E., Rite, S., Santiago, M., Romo, M., Samper, S., Cebollada, A., Oteo-Iglesias, J., Rezusta, A., 2021. Successful control of *Serratia marcescens* outbreak in a neonatal unit of a tertiary-care hospital in Spain. Enferm Infecc Microbiol Clin (Engl Ed) S0213–005X(21)00186-5. 10.1016/j.eimc.2021.05.003

32. Murray, C.J., Ikuta, K.S., Sharara, F., Swetschinski, L., Aguilar, G.R., Gray, A., Han, C., Bisignano, C., Rao, P., Wool, E., Johnson, S.C., Browne, A.J., Chipeta, M.G., Fell, F., Hackett, S., Haines- Woodhouse, G., Hamadani, B.H.K., Kumaran, E.A.P., McManigal, B., Agarwal, R., Akech, S., Albertson, S., Amuasi, J., Andrews, J., Aravkin, A., Ashley, E., Bailey, F., Baker, S., Basnyat, B., Bekker, A., Bender, R., Bethou, A., Bielicki, J., Boonkasidecha, S., Bukosia, J., Carvalheiro, C., Castañeda-Orjuela, C., Chansamouth, V., Chaurasia, S., Chiurchiù, S., Chowdhury, F., Cook, A.J., Cooper, B., Cressey, T.R., Criollo-Mora, E., Cunningham, M., Darboe, S., Day, N.P.J., Luca, M.D., Dokova, K., Dramowski, A., Dunachie, S.J., Eckmanns, T., Eibach, D., Emami, A., Feasey, N., Fisher- Pearson, N., Forrest, K., Garrett, D., Gastmeier, P., Giref, A.Z., Greer, R.C., Gupta, V., Haller, S., Haselbeck, A., Hay, S.I., Holm, M., Hopkins, S., Iregbu, K.C., Jacobs, J., Jarovsky, D., Javanmardi, F., Khorana, M., Kissoon, N., Kobeissi, E., Kostyanev, T., Krapp, F., Krumkamp, R., Kumar, A., Kyu, H.H., Lim, C., Limmathurotsakul, D., Loftus, M.J., Lunn, M., Ma, J., Mturi, N., Munera-Huertas, T., Musicha, P., Mussi-Pinhata, M.M., Nakamura, T., Nanavati, R., Nangia, S., Newton, P., Ngoun, C., Novotney, A., Nwakanma, D., Obiero, C.W., Olivas-Martinez, A., Olliaro, P., Ooko, E., Ortiz-Brizuela, E., Peleg, A.Y., Perrone, C., Plakkal, N., Ponce-de-Leon, A., Raad, M., Ramdin, T., Riddell, A., Roberts, T., Robotham, J.V., Roca, A., Rudd, K.E., Russell, N., Schnall, J., Scott, J.A.G., Shivamallappa, M., Sifuentes-Osornio, J., Steenkeste, N., Stewardson, A.J., Stoeva, T., Tasak, N., Thaiprakong, A., Thwaites, G., Turner, C., Turner, P., Doorn, H.R. van Velaphi, S., Vongpradith, A., Vu, H., Walsh, T., Waner, S., Wangrangsimakul, T., Wozniak, T., Zheng, P., Sartorius, B., Lopez, A.D., Stergachis, A., Moore, C., Dolecek, C., Naghavi, M., 2022. Global burden of bacterial antimicrobial resistance in 2019: a systematic analysis. The Lancet 399, 629–655. 10.1016/S0140-6736(21)02724-0

33. Pirzadian, J., Harteveld, S.P., Ramdutt, S.N., van Wamel, W.J.B., Klaassen, C.H.W., Vos, M.C., Severin, J.A., 2020. Novel use of culturomics to identify the microbiota in hospital sink drains with and without persistent VIM-positive *Pseudomonas aeruginosa*. Sci Rep 10, 17052. 10.1038/s41598-020-73650-8

34. Ramos-Castaneda, J.A., Faron, M.L., Hyke, J., Bell-Key, D., Buchan, B.W., Nanchal, R., Pintar, P., Graham, M.B., Huerta, S., Munoz-Price, L.S., 2020. How frequently should sink drains be disinfected? Infect Control Hosp Epidemiol 41, 358–360. 10.1017/ice.2019.316

35. Reissier, S., Cattoir, V., 2021. Streptogramins for the treatment of infections caused by Gram-positive pathogens. Expert Rev Anti Infect Ther 19, 587–599. 10.1080/14787210.2021.1834851

36. Rodriguez-Mozaz, S., Chamorro, S., Marti, E., Huerta, B., Gros, M., Sanchez-Melsio, A., Borrego, C.M., Barcelo, D., Balcazar, J.L., 2015. Occurrence of antibiotics and antibiotic resistance genes in hospital and urban wastewaters and their impact on the receiving river. Water Research Volume 69, 234--242.

37. Schwering, M., Song, J., Louie, M., Turner, R.J., Ceri, H., 2013. Multi-species biofilms defined from drinking water microorganisms provide increased protection against chlorine disinfection. Biofouling 29, 917–928. 10.1080/08927014.2013.816298

38. Sivalingam, P., Poté, J., Prabakar, K., 2020. Extracellular DNA (eDNA): Neglected and Potential Sources of Antibiotic Resistant Genes (ARGs) in the Aquatic Environments. Pathogens 9, 874. 10.3390/pathogens9110874

39. Snell, L.B., Prossomariti, D., Alcolea-Medina, A., Sasson, M., Dibbens, M., Al-Yaakoubi, N., Humayun, G., Charalampous, T., Alder, C., Ward, D., Maldonado-Barrueco, A., Abadioru, O., Batra, R., Nebbia, G., Otter, J.A., Edgeworth, J.D., Goldenberg, S.D., 2024. The drainome: longitudinal metagenomic characterisation of wastewater from hospital ward sinks to characterize the microbiome and resistome and assess the effects of decontamination interventions. Journal of Hospital Infection. 10.1016/j.jhin.2024.06.005

40. Suleyman, G., Alangaden, G., Bardossy, A.C., 2018. The Role of Environmental Contamination in the Transmission of Nosocomial Pathogens and Healthcare-Associated Infections. Curr Infect Dis Rep 20, 12. 10.1007/s11908-018-0620-2

41. van der Veen, S., Abee, T., 2011. Mixed species biofilms of *Listeria monocytogenes* and *Lactobacillus plantarum* show enhanced resistance to benzalkonium chloride and peracetic acid. Int J Food Microbiol 144, 421–431. 10.1016/j.ijfoodmicro.2010.10.029

42. van der Zwet, W.C., Nijsen, I.E.J., Jamin, C., van Alphen, L.B., von Wintersdorff, C.J.H., Demandt, A.M.P., Savelkoul, P.H.M., 2022. Role of the environment in transmission of Gram-negative bacteria in two consecutive outbreaks in a haematology-oncology department. Infect Prev Pract 4, 100209. 10.1016/j.infpip.2022.100209

43. Volling, C., Ahangari, N., Bartoszko, J.J., Coleman, B.L., Garcia-Jeldes, F., Jamal, A.J., Johnstone, J., Kandel, C., Kohler, P., Maltezou, H.C., Maze dit Mieusement, L., McKenzie, N., Mertz, D., Monod, A., Saeed, S., Shea, B., Stuart, R.L., Thomas, S., Uleryk, E., McGeer, A., 2021. Are Sink Drainage Systems a Reservoir for Hospital-Acquired Gammaproteobacteria Colonization and Infection? A Systematic Review. Open Forum Infectious Diseases 8, ofaa590. 10.1093/ofid/ofaa590

44. Wingender, J., Flemming, H.C., 2011. Biofilms in drinking water and their role as reservoir for pathogens. Int.J.Hyg.Environ.Health 214, 417–423. 10.1016/j.ijheh.2011.05.009

