## Supplementary Figures for "The hospital sink drain biofilm resistome is independent of the corresponding microbiota, the environment and disinfection measures"

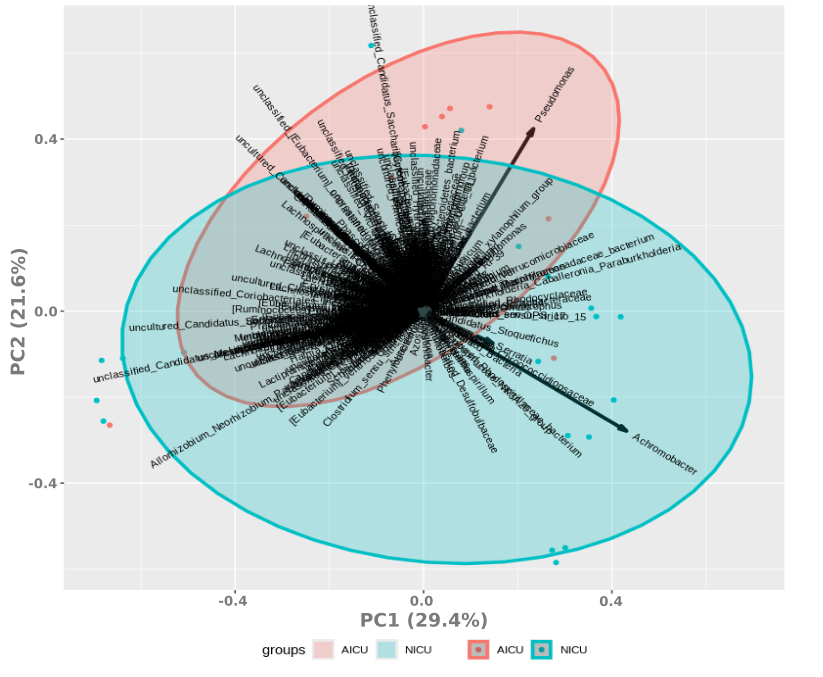

**Supplementary Fig. 1**: **Principal Coordinate Analysis (PCA) visualizing the microbiota of sink-drain biofilms at the NICU (blue) and AICU (red) determined by full-length 16S rRNA**. Depicted taxa are on genus level. Data were normalized using logarithmic and Hellinger’s transformations.

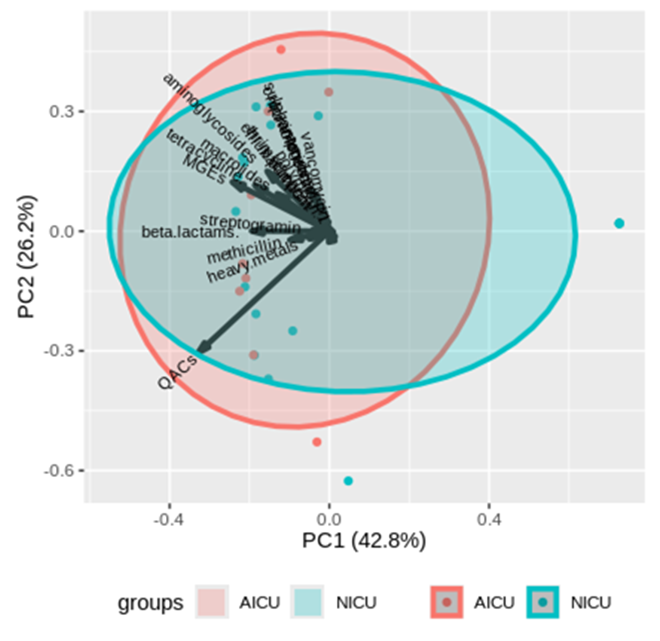

**Supplementary Fig. 2**: **Principal Coordinate Analysis (PCA) visualizing the resistome of NICU (blue) and AICU (red) sink-drain biofilms.** Normalized abundance in sink-drain biofilms samples of individual genes was accumulated per resistance gene class. Data were normalized using logarithmic and Hellinger’s transformations.

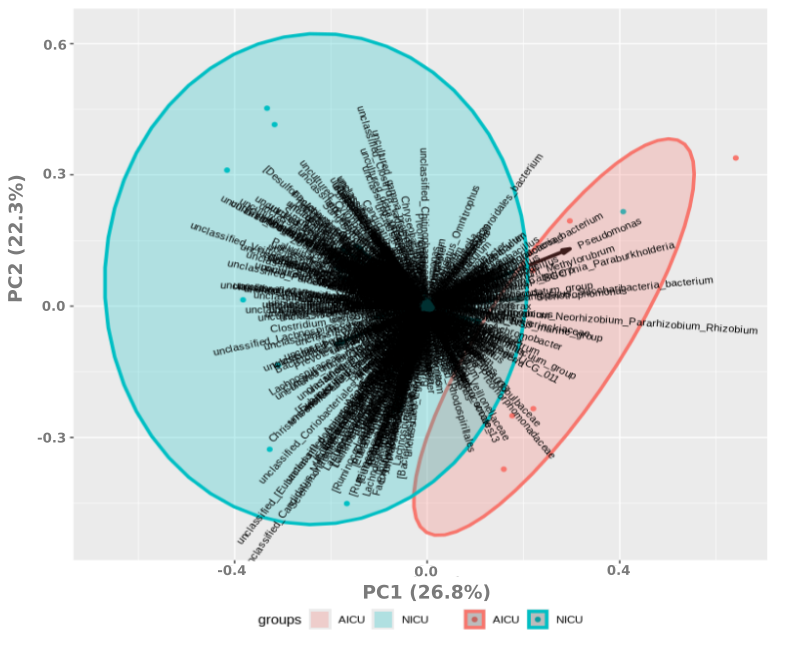

**Supplementary Fig. 3**: **Principal Coordinate Analysis (PCA)** **visualizing the microbiota of WW and WW biofilms at the NICU (blue) and AICU (red) determined by full-length 16S rRNA.** Depicted taxa are on genus level. Data were normalized using logarithmic and Hellinger’s transformations.

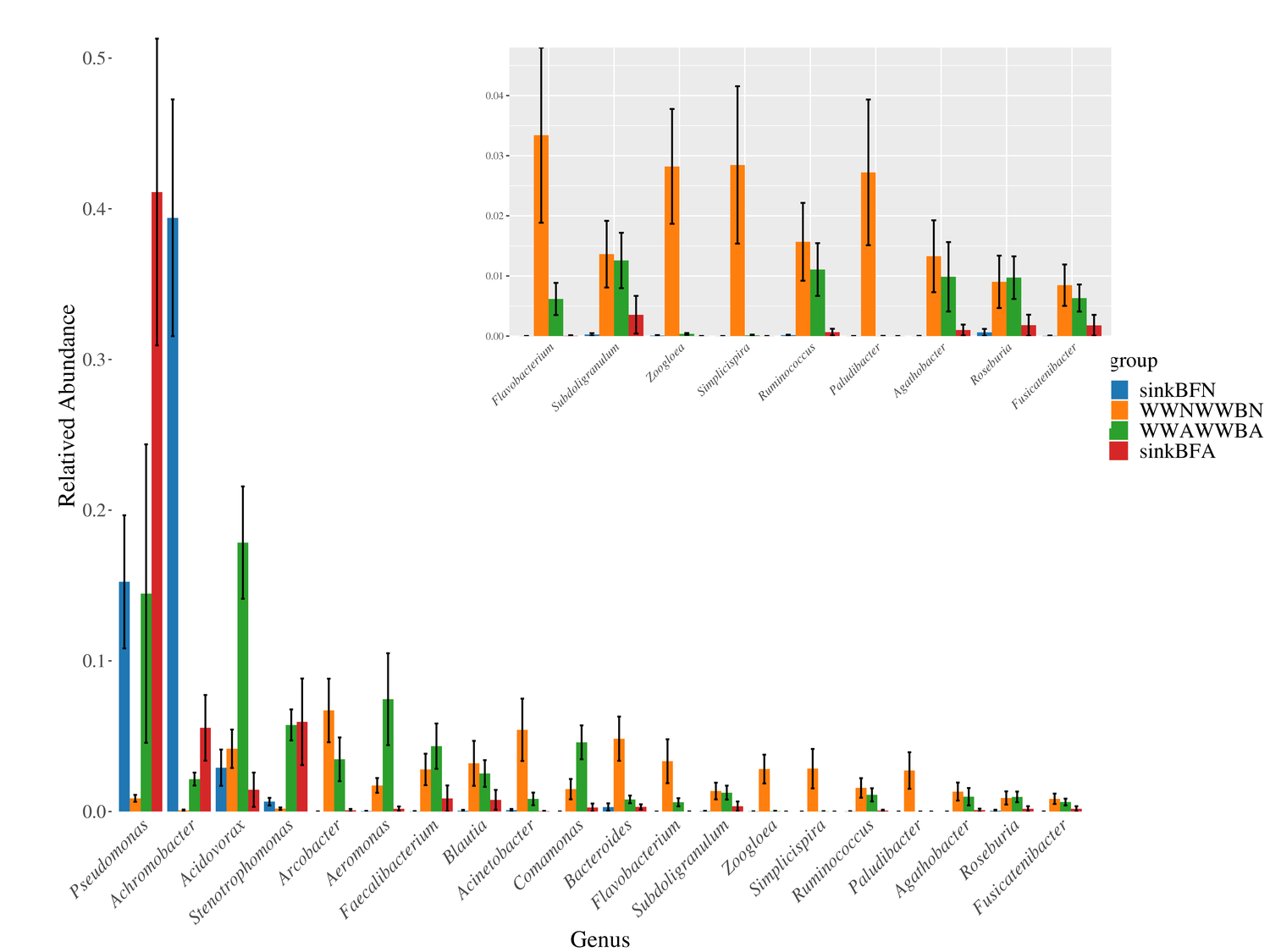

**Supplementary Fig. 4:** **The most abundant genera in sink-drain biofilms and NICU or AICU WW.** Relative abundance was plotted only for the most abundance bacterial genus in sink-drain or wastewater samples. Anova was performed for each sample location (AICU or NICU) in both types of samples.

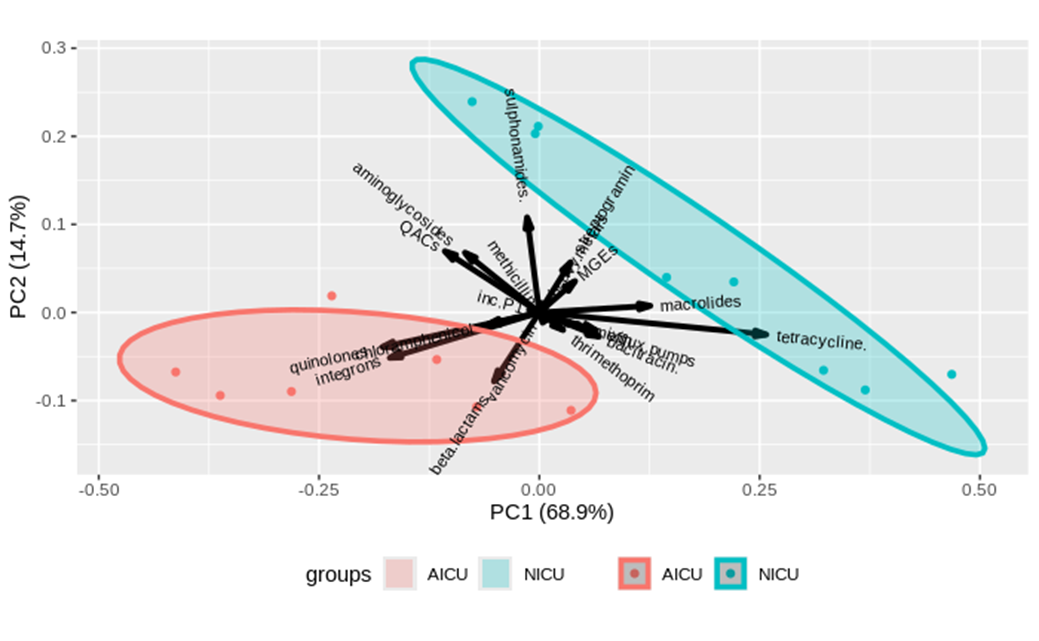

**Supplementary Fig. 5:** **Principal Coordinate Analysis (PCA) visualizing the resistome of NICU WW and WW biofilm (blue) and AICU WW and WW biofilm (red).** Normalized abundance in wastewater samples of individual genes was accumulated per resistance gene class. Data were normalized using logarithmic and Hellinger’s transformations.
